# Evaluation of accelerated whole-body diffusion weighted imaging with deep learning reconstruction in patients with metastatic prostate cancer: assessment of image quality and ADC estimates

**DOI:** 10.64898/2026.09.17.26362983

**Authors:** Evanthia Kousi, Mihaela Rata, Dow-Mu Koh, Christina Messiou, Nina Tunariu, Georgina Hopkinson, Emily Evans, Omar Darwish, Elisabeth Weiland, Jessica M Winfield

## Abstract

**Objectives:** To evaluate the impact of an accelerated whole-body diffusion-weighted MRI (WB-DWI) protocol using deep learning (DL) reconstruction on image quality and apparent diffusion coefficient (ADC) quantification in patients with metastatic prostate cancer.

**Methods:** This single-centre prospective study involved two patient cohorts undergoing standard-of-care WB-MRI at 1.5T. The DL WB-DWI acquisition used fewer signal averages and higher parallel imaging acceleration, reconstructed with a research application DL package based on a variational network. In Cohort 1 (*n* = 10), qualitative assessment of image quality was performed across four anatomical stations using a 4-point Likert scale by two radiologists, blinded to the DWI protocol. In Cohort 2 (n = 20), ADC values were evaluated in hypercellular focal bone metastases (as determined by a radiologist) and compared between the standard and accelerated DL WB-DWI protocols. Image quality scores were analysed using Wilcoxon signed-rank tests (Bonferroni-corrected), and ADC agreement was evaluated via Bland–Altman analysis.

**Results:** Accelerated DL WB-DWI reduced WB-DWI acquisition time by 37% without compromising image quality. Radiologists consistently rated both protocols as good-to-excellent across all image quality metrics. Median ADC estimates in lesions showed no significant difference between protocols; interquartile range of ADC estimates also showed no significant difference.

**Conclusions:** Accelerated DL WB-DWI enables 37% scan time reduction without degrading image quality or affecting ADC estimates of lesions in patients with metastatic prostate cancer.

**Advances in knowledge:** DL-based reconstruction enables faster WB-DWI acquisition without compromising image quality or ADC quantification, supporting clinical implementation in metastatic prostate cancer staging and therapy assessment.

## Introduction

Prostate cancer is one of the most prevalent malignancies in men and remains a significant contributor to cancer-related mortality worldwide [1]. Accurate imaging of bone metastases from prostate cancer is critical for detection, staging, monitoring disease progression and assessment of therapeutic response. Whole-body diffusion-weighted imaging (WB-DWI) is an established imaging technique, offering functional insights into tissue microstructure and tumour burden [2]. The apparent diffusion coefficient (ADC) derived from DWI is considered a marker of tumour cellularity and cell membrane integrity with multiple studies showing a correlation between ADC increase and the reduction in tumour cellularity following successful therapy [3, 4, 5].

However, the clinical adoption of WB-DWI is hampered by several challenges, most notably the long acquisition times due to the high number of signal averages acquired to mitigate the inherent low signal-to-noise ratio (SNR) of the DWI technique. Long acquisition times lead to difficulty in scheduling examinations in busy radiology departments and lengthy examinations can be difficult for patients to tolerate which may in turn lead to increased motion and a reduction in image quality.

The introduction of deep learning (DL)-based reconstruction for several DWI sequences and body regions has shown great potential to accelerate MR imaging acquisitions while preserving and/or enhancing the diagnostic image quality [6, 7]. Prior studies have applied these methods to various anatomical sites such as brain [8], pelvis [6, 9], abdomen [10, 11], breast [12, 13], spine [7], and whole-body [14] with promising results. Recently, Ponsiglione et al. [14] evaluated the clinical feasibility of an accelerated DL WB-DWI and showed improved image quality compared to conventional sequences. However, these studies often focus on general image quality assessments and acquisition time reduction, with limited emphasis on cancer type-specific quantitative evaluations or ADC estimates.

The aim of this study is to evaluate the performance of a research DL-based reconstruction in WB-DWI, by comparing image quality and ADC estimates in accelerated DL WB-DWI versus standard WB-DWI acquisitions in patients with metastatic prostate cancer.

## Methods

### Patients

This prospective single-centre study was approved by a national research ethics committee (NCT05118555, clinicaltrials.gov). All patients provided verbal consent for the acquisition of additional research imaging data at the end of their routine clinical examinations.

### MRI Acquisition

Two cohorts of patients were scanned on a 1.5T scanner (MAGNETOM Sola, Siemens Healthineers, Forchheim, Germany) between April 2024 and January 2025. All patients underwent routine clinical WB-MRI (including WB-DWI and Dixon sequences) using five anatomical stations (head to thighs). The research sequences were performed at the end of the examination. Details of the two cohorts and their specific imaging protocols are provided in later sections.

The study protocol was based on the standard WB-DWI protocol at our institution (hereafter referred to as protocol A), which employed the DWI pulse sequence supplied by the scanner manufacturer. An accelerated DL WB-DWI (using the research prototype sequence) was investigated (hereafter referred to as protocol B). For completeness, a replica of protocol A was created using the research prototype sequence (see supplementary material).

Image quality and ADC estimates were compared between the standard DWI (protocol A) and the accelerated DL DWI protocol (protocol B). Protocol B was implemented using fewer signal averages and higher parallel imaging acceleration factor. The acquisition parameters of all protocols are summarised in Table 1.

**Table 1:**
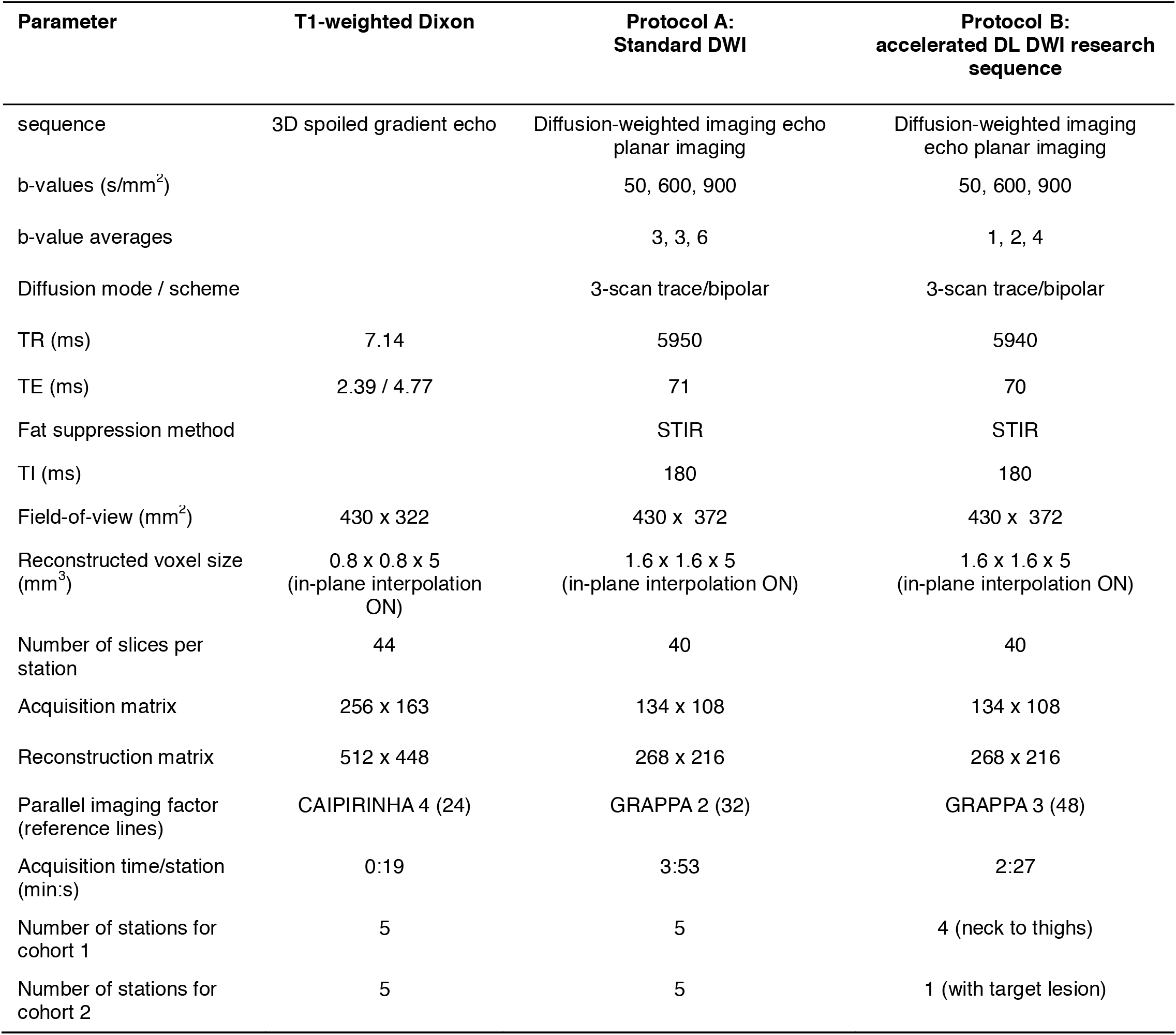
WB-DWI and WB-T1-weighted Dixon acquisition parameters.

### DL-Based DWI reconstruction

DL-DWI data were reconstructed inline using a prototype DL-based variational network [15], trained on approximately 500,000 single-shot DW images from healthy volunteers acquired on 1.5T and 3T scanners (MAGNETOM, Siemens Healthineers, Forchheim, Germany) [10]. The network alternated between data consistency and learned regularization steps, incorporating precalculated coil sensitivity maps and a hierarchical convolutional neural network (CNN)-based module. Training was performed offline in PyTorch on an NVIDIA GPU Cluster (Tesla V100-SXM2), and the network was frozen and deployed on the scanner using a C++ inference framework. Standard post-processing (averaging, trace-weighting, and ADC calculation) was applied identically to standard DWI.

### Image quality assessments (Cohort 1)

In Cohort 1 (n = 10, mean age 70 ± 7 years, BMI = 29 ± 6 kg/m^2^), image quality metrics were assessed across a 4-station WB-DWI acquisition (from neck to thighs, the head station was excluded) and compared between protocol A (standard DWI) and protocol B (accelerated DL DWI research sequence). All readings were performed on a PACS workstation (IDS7, Sectra, Sweden). The added examination time for cohort 1 was 12 min and 15 s for four stations. Two radiologists with more than□20 years of experience, blinded to the WB-DWI protocol, independently scored the images using a 4-point Likert scale (1 = unacceptable/non-diagnostic, 2 = adequate, 3 = good, 4 = excellent). The scoring assessed the overall image quality for b900, ADC, and b900 maximum intensity projection (MIP) images, and b900 lesion conspicuity across all stations. In addition, scoring was performed for each station separately to assess SNR, sharpness and artifacts on b900 images and artifacts on ADC images.

### ADC estimates assessments (Cohort 2)

In cohort 2 (n = 20, mean age 75 ± 9 years, BMI = 28 ± 5 kg/m^2^) images were assessed using both protocol A (standard DWI) and protocol B (accelerated DL DWI research sequence) in hypercellular focal bone metastases (size ≥ 1cm), as determined by a radiologist, from a single station. The lesions were determined hypercellular based on following criteria: hyperintense on b900, low ADC (500-900 mm2/s) and low relative fat fraction (rFF less than 20%) based on previously published data [16].

The ADC estimates from protocol B (accelerated DL DWI research sequence) were compared with the ADC estimates from protocol A (standard DWI).

Focal lesions were manually delineated on all slices encompassing the lesion on standard b900 images (Protocol A), and the volumes of interest (VOIs) were transferred to the corresponding ADC maps from all protocols for each patient (IDL ADEPT, ICR, UK). All VOIs were visually verified on fat fraction (FF) maps to avoid fully treated tumours (Figure 1). FF maps were generated using Dixon acquisitions (Table 1) as part of the standard WB-MRI protocol.

**Figure 1:**
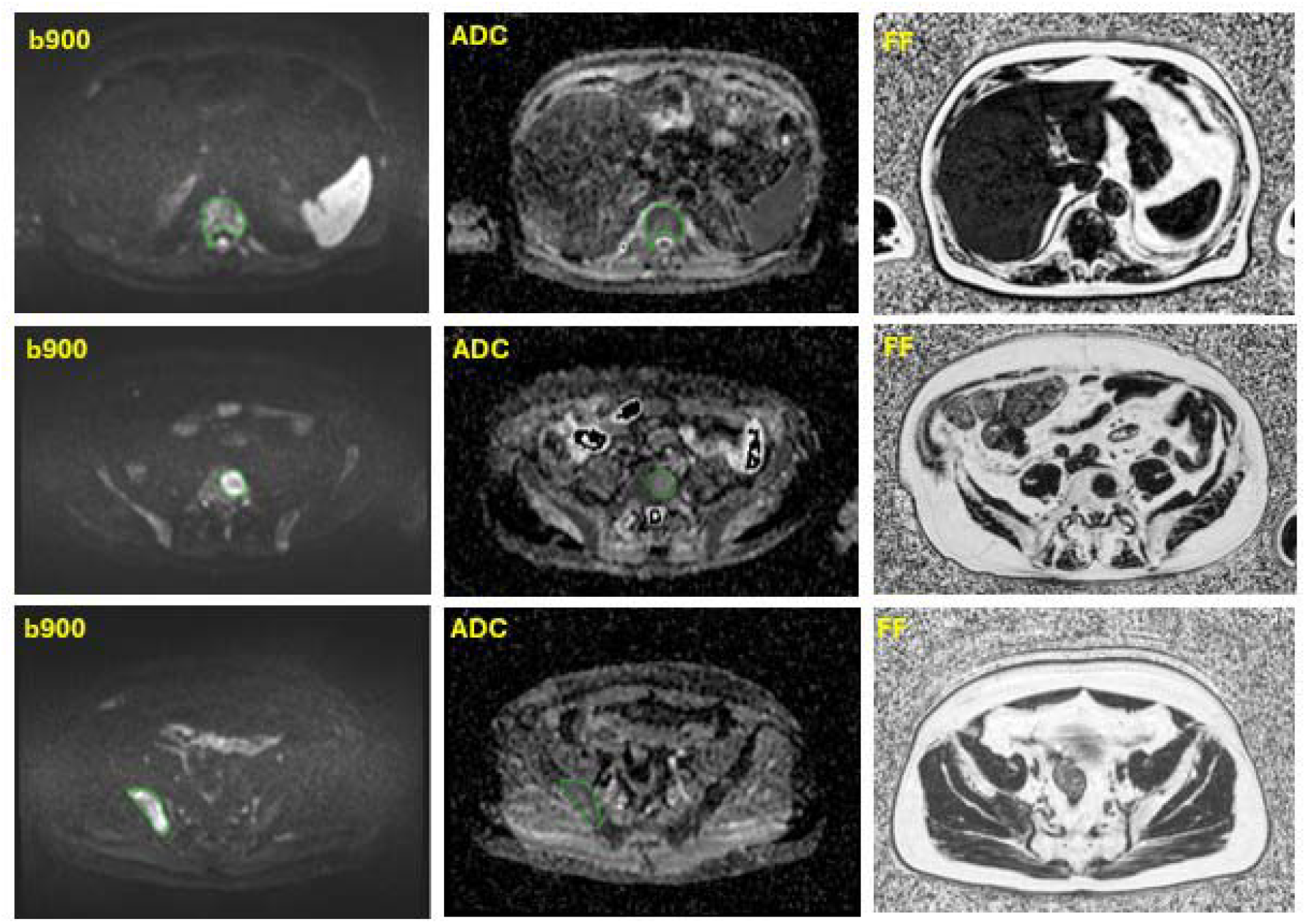
Examples of tumour delineation on standard WB-DWI b900 images for three patients with lesions in T11 (top row), L5 (middle row) and right iliac bone (bottom row). All ROIs (show in green) were viewed alongside the corresponding FF maps to avoid areas of fully treated tumour. All ADC maps are displayed at the same window width and level.

The median and interquartile range (IQR) of ADC estimates were calculated from all fitted voxels in the VOI for each patient.

### Statistical analysis

#### Image quality (Cohort 1)

For each reader, the differences between the median image quality Likert scores for protocol A (standard DWI) and protocol B (accelerated DL DWI research sequence) were evaluated using the paired 2-sided Wilcoxon signed-rank test (MATLAB, R2024b, MathWorks). For each reader, there were 20 statistical comparisons (4 separate comparisons across the 4-stations plus 16 separate comparisons for the separate stations considered), and therefore the Bonferroni corrected significance threshold was set at p = 0.05/20 = 0.0025.

#### ADC estimates (Cohort 2)

The mean and standard deviation of cohort 2 median ADC estimates for each DWI protocol (protocol A and protocol B) are reported based on measurements from focal lesions. Bland-Altman analysis and the paired 2-sided Wilcoxon signed-rank test (p = 0.05) were used to assess whether median and IQR ADC estimates differed significantly between protocol A (standard DWI) and protocol B (accelerated DL DWI research sequence).

## Results

### Image Quality assessments (Cohort 1)

The acquisition time of a station was reduced by 37% when using protocol B (2:27 min per station) compared to protocol A (3:53 min per station) which corresponds to a total reduction in acquisition time of 7 minutes and 10 seconds for a 5 station WB-DWI acquisition.

Image quality was rated similarly between protocol A (standard DWI) and protocol B (accelerated DL DWI research sequence), with no statistically significant differences across any qualitative metric (p > 0.0025 for all comparisons). Most ratings were “good” (3), with some rated as “excellent” (4), across b900, ADC, and b900 MIP images for both readers, as illustrated by the boxplots in figure 2.

**Figure 2:**
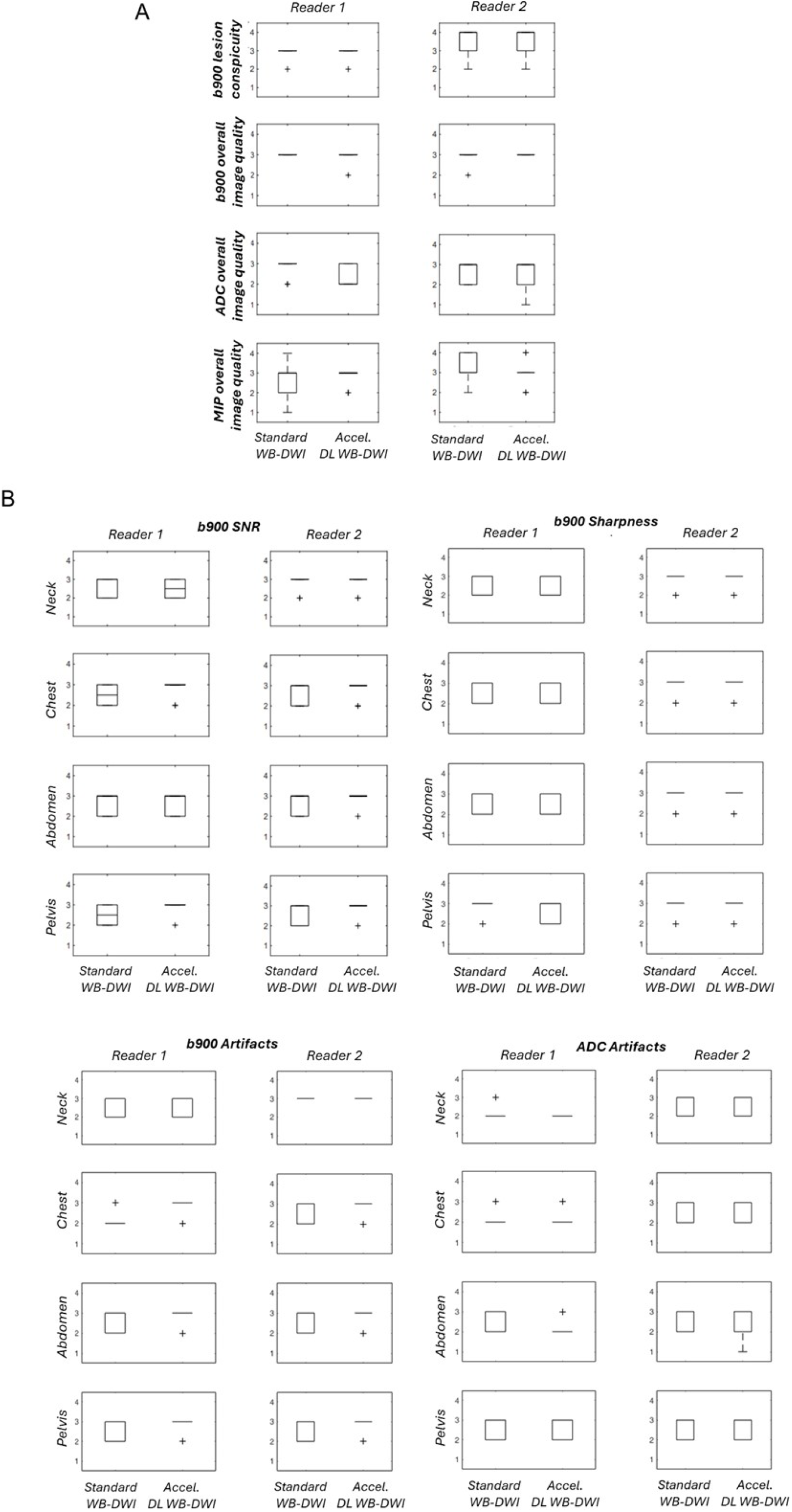
Boxplots illustrate the Likert scale scores (1= unacceptable/non-diagnostic, 2= adequate, 3= good, 4= excellent) of image quality metrics in standard WB-DWI (protocol A) and accelerated DL WB-DWI (protocol B) for reader 1 and reader 2. Lesion conspicuity in b900 images and the overall image quality of b900, ADC and b900 MIP images were assessed across all four stations (A), while b900 image SNR, sharpness and artifacts as well as ADC artifacts were assessed for each station separately (B). Lower artifact scores indicate more artifacts. Bonferroni corrected p-value of significance p = 0.0025.

Examples of b900, ADC and b900 MIP images acquired with protocol A (standard WB-DWI) and protocol B (accelerated DL WB-DWI) are shown in figure 3.

**Figure 3:**
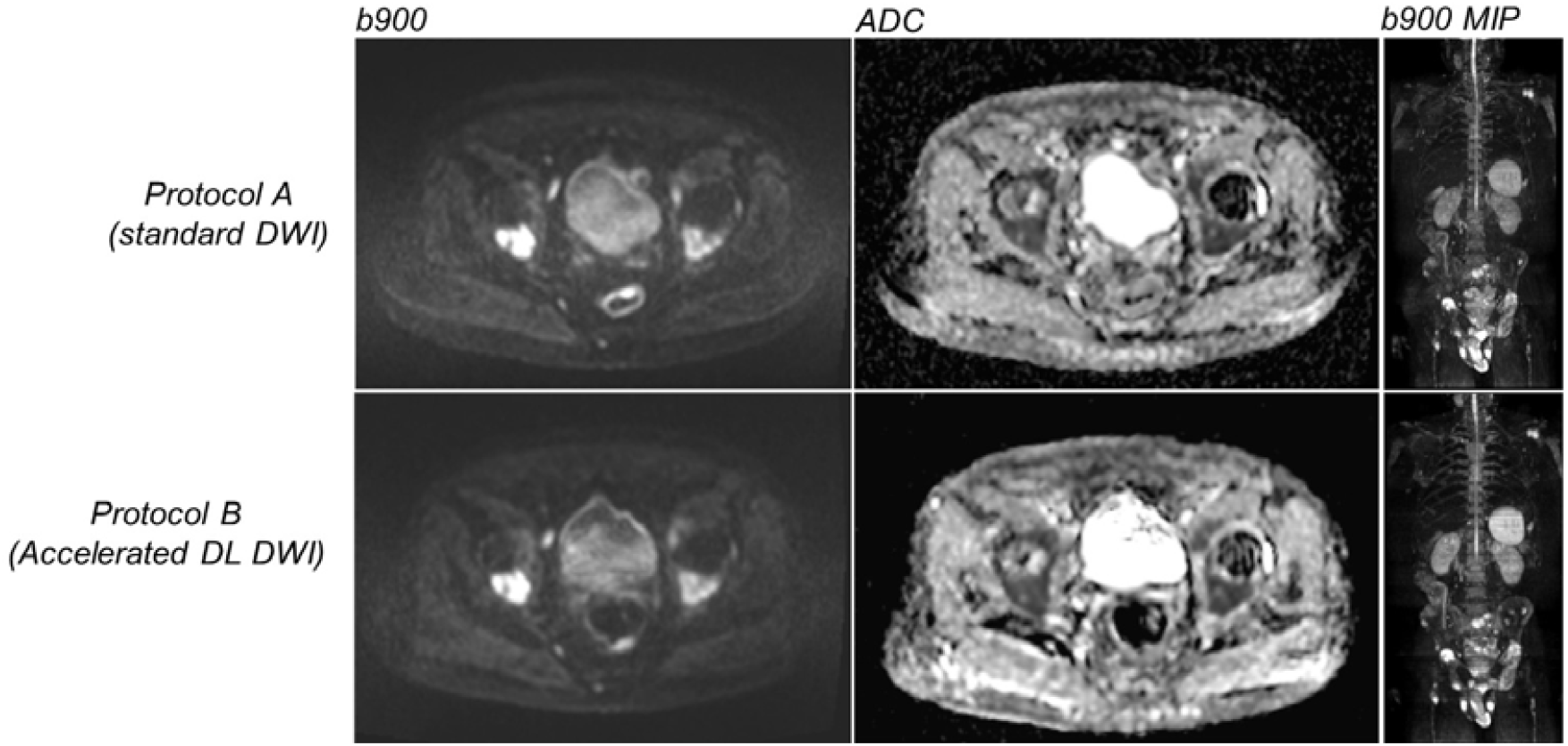
Examples of b900 and ADC images of the pelvis as well as b900 MIP images from an 82-year-old patient in cohort 1 with bone metastatic disease, acquired with standard WB-DWI (top row) and accelerated DL WB-DWI (bottom row). The ADC maps are displayed at the same window width and level.

### ADC estimates assessments (Cohort 2)

The mean lesion volume in Cohort 2 was 16.7 mm^3^ (range: 2.4–95 mm^3^). The cohort mean and standard deviations of the median lesion ADC values from protocol A (standard DWI) and protocol B (accelerated DL DWI research sequence) are shown in table 2.

**Table 2.**
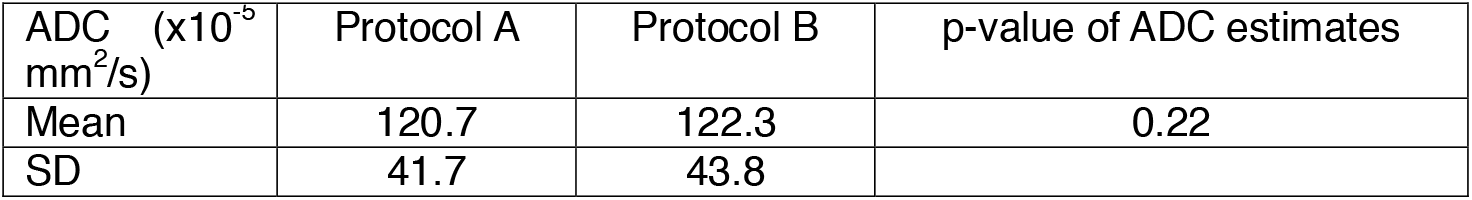
Summary of ADC estimates (mean and standard deviation, SD) using protocols A (standard DWI) and B (accelerated DL DWI research sequence) across all 20 lesions.

There were no statistically significant differences between the ADC values estimated from protocol B (accelerated DL DWI research sequence) and protocol A (standard DWI) (p = 0.22). Similarly, the IQR of ADC values was not significantly different (p = 0.89). Bland–Altman analysis of median ADC (figures 4a) and IQR (figure 4b) demonstrated strong agreement, with minimal bias (−1.6 and 0.11) and no clear dependence of the difference on the mean values. The 95% limits of agreement were −9.9 to 6.7 for median ADC and −10.1 to 10.3 for IQR.

**Figure 4:**
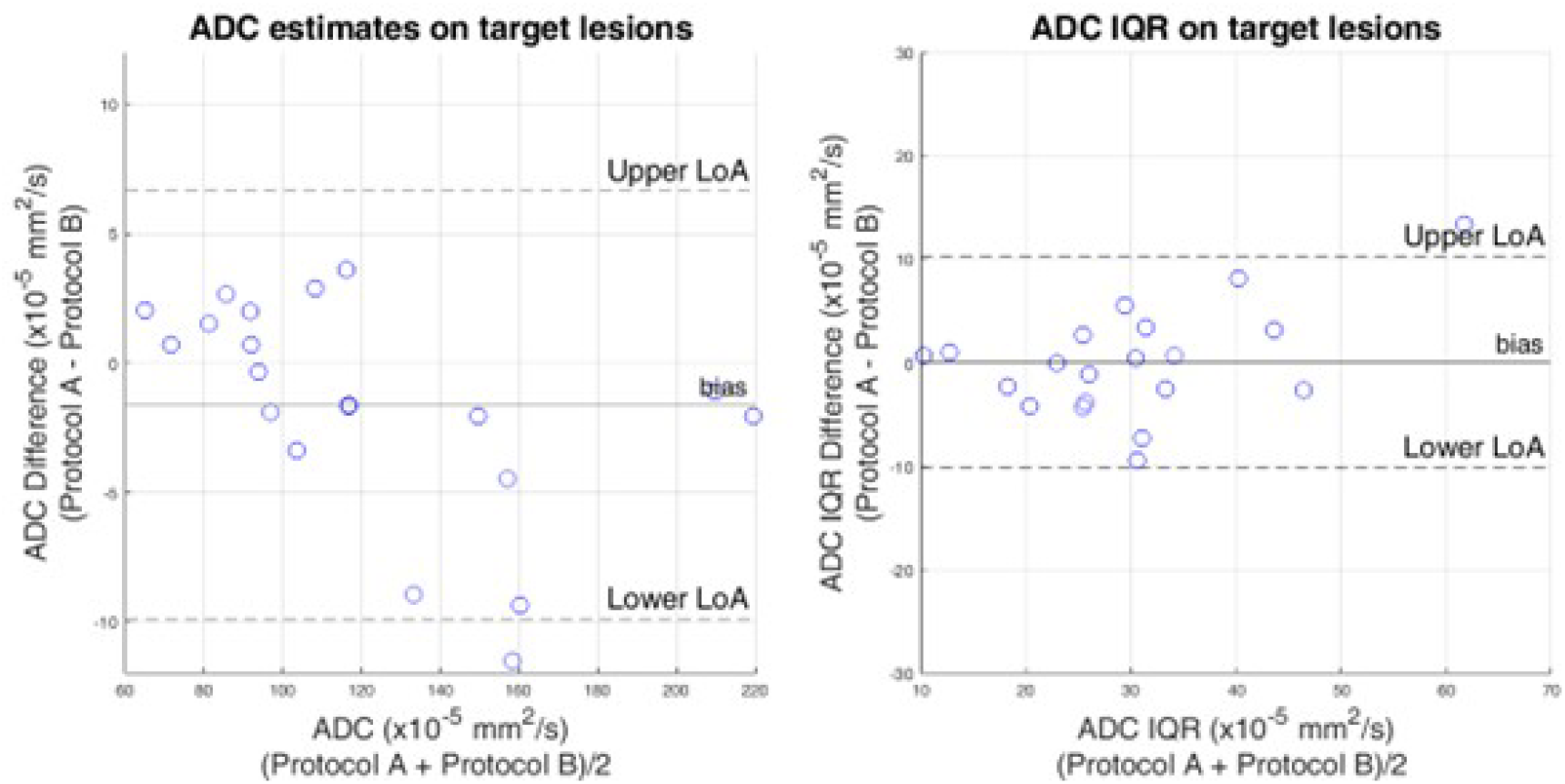
Bland–Altman plots of median (left) and IQR ADC (right) measurements demonstrating agreement between the two acquisition protocols in target bone lesions in Cohort 2. Two data points have nearly identical median ADC values and overlap, appearing as a single point on the left plot.

## Discussion

The good image quality in accelerated DL WB-DWI and the absence of differences in ADC estimates shows that faster WB-DWI scans can be achieved without compromising diagnostic quality and quantitative accuracy.

The qualitative assessments in our 10-patient cohort demonstrated that an accelerated DWI protocol with DL-based reconstruction can reduce acquisition time by 7min and 10s (or 37%) for a 5-station WB-DWI examination whilst maintaining image quality.

The benefit of DL-based denoising algorithms in image quality for WB-DWI has been previously demonstrated by Zormpas-Petridis et al. [17] who developed a U-Net deep learning algorithm for WB-DWI and demonstrated reduced imaging times (from 25-30 minutes to approximately 5 minutes for 4-5 stations) while also improving the image quality of high b-value images and ADC maps. Their approach included an additional processing step after image reconstruction to recover noisy DWI datasets acquired with a reduced number of averages. In another recent WB study, Ponsiglione et al. showed that an accelerated DL DWI protocol along with other protocol changes, significantly improved image quality in both normal tissues and cancer lesions when compared with a standard WB-DWI sequence. The study, which included a heterogeneous cohort of patients with myeloma as well as advanced prostate and breast cancers, also reported reductions in acquisition time of more than 50% [14].

The impact of accelerated DL DWI has also been investigated in the pelvis by Hermann et al. [6] in a cohort of patients with various pelvic pathologies, including neoplasms. In their study, scan acceleration was simulated by retrospectively applying a DL algorithm to raw data derived from a reduced number of the originally acquired b-value averages. The authors reported that the DL-based approach preserved both the overall image quality of DWI and diagnostic confidence, while enabling a 40% reduction in acquisition time. A similar approach was followed by Johnson et al. in prostate retrospectively generating three- and four-fold accelerated DWI datasets [18]. The authors found no significant differences in the overall image quality, artifacts and anatomical clarity between a standard and a DL DWI sequence when comparing both low and high b-value images.

Other studies retrospectively evaluated the use of a DL algorithm to enhance image quality in spine and prostate DWI, without focusing on acceleration and thus utilising identical acquisition parameters between the DWI protocols [7, 9]. In a large whole-spine study Kim et al. found improved overall image quality and diagnostic confidence using DL-reconstructed DWI whereas the detection rates of malignant disease were not significantly different between the standard and the DL DWI protocols [7]. Similarly, Ueda et al, showed improved SNR and contrast-to-noise ratio (CNR) for prostate images acquired with DL DWI compared with a standard DWI sequence [9].

In our cohort of 20 patients, no statistically significant differences were observed in ADC values within bone marrow lesions when comparing accelerated DL DWI with standard DWI. These findings agree with previous studies focusing on the prostate, breast and whole-spine showing no significant differences in ADC between accelerated DL DWI and standard DWI protocols [7, 9, 13]. The consistency of ADC measurements observed in our study, as well as in previous reports [7, 9, 13], may be attributed to the use of largely comparable sequence parameters. In these studies, the DWI protocols primarily differed in the number of signal averages while maintaining other technical parameters such as diffusion mode, diffusion scheme and fat suppression method. Ponsiglione et al assessed the influence of an accelerated DL DWI sequence in WB with more extensive parameter changes and found significantly higher ADC values for brain and psoas muscle but not for bone metastases compared with a standard protocol [14]. Nevetherless, the authors focus was to assess the net benefit of an optimised DL-based DWI sequence, acknowledging the technical variability between the sequences involved and the impact it may have on their results.

Other studies focusing on abdominal DWI have reported conflicting results on the impact of DL-based reconstructions on ADC estimates. Bae et al. [11] observed significantly lower ADC values in both normal liver tissue and focal hepatic lesions, while Afat et al. [10] reported significantly greater ADC values across several abdominal tissues for accelerated DL DWI compared with standard DWI protocols.

While the design of the presented study allowed for the comparison of image quality and ADC estimates between standard and accelerated DWI protocols in a specialised patient cohort, the relatively small cohort size and single-centre study, may limit the generalisability of our findings. However, the study included patients with metastatic prostate cancer undergoing WB-MRI examinations at any treatment stage and is therefore representative of this WB-MRI patient population. Larger multicentre studies will be needed to validate these results further and explore consistency across different scanner platforms, DL implementations and patient populations.

In conclusion, our study confirms that accelerated DL WB-DWI protocols have the potential to achieve substantial scan time reductions without sacrificing diagnostic image quality or ADC quantification in metastatic prostate cancer patients. This represents a promising step towards more efficient WB-MRI workflows, particularly for oncologic patients requiring serial imaging.

## Data Availability

All data produced in the present study are available upon reasonable request to the authors

