## Supplementary material for "Evaluation of accelerated whole-body diffusion weighted imaging with deep learning reconstruction in patients with metastatic prostate cancer: assessment of image quality and ADC estimates"

A replica of the standard DWI was created using the research prototype sequence (non-accelerated research DWI). The non-accelerated research DWI sequence, served as the basis for the accelerated DL DWI sequence (protocol B). The acquisition parameters of the non-accelerated research DWI are shown in the supplementary table 1.

| **Supplementary table 1**: acquisition parameters of the standard DWI and replica sequence using the research sequence (non-accelerated DWI research sequence. | | |
| --- | --- | --- |
| **Parameter** | **Standard DWI** | **Non-accelerated research DWI** |
| sequence | Diffusion-weighted imaging echo planar imaging | Diffusion-weighted imaging echo planar imaging |
| b-values (s/mm^2^) | 50, 600, 900 | 50, 600, 900 |
| b-value averages | 3, 3, 6 | 3, 3, 6 |
| Diffusion mode / scheme | 3-scan trace/bipolar | 3-scan trace/bipolar |
| TR (ms) | 5950 | 6290 |
| TE (ms) | 71 | 74 |
| Fat suppression method | STIR | STIR |
| TI (ms) | 180 | 180 |
| Field-of-view (mm^2^) | 430 x 372 | 430 x 372 |
| Reconstructed voxel size (mm^3^) | 1.6 x 1.6 x 5  (in-plane interpolation ON) | 1.6 x 1.6 x 5  (in-plane interpolation ON) |
| Number of slices per station | 40 | 40 |
| Acquisition matrix | 134 x 108 | 134 x 108 |
| Reconstruction matrix | 268 x 216 | 268 x 216 |
| Parallel imaging factor (reference lines) | GRAPPA 2 (32) | GRAPPA 2 (32) |
| Acquisition time/station (min:s) | 3:53 | 4:06 |
| Number of stations for cohort 1 | 5 | N/A |
| Number of stations for cohort 2 | 5 | 1 (with target lesion) |

For completeness the ADC estimates in cohort 2 were compared between the standard DWI and the non-accelerated research DWI to assess agreement. The ADC estimates for each protocol across all 20 lesions are shown in the supplementary table 2.

There were no statistically significant differences between the ADC estimates from the standard DWI and the non-accelerated research DWI (p = 0.11) and Bland-Altman analysis demonstrated good agreement between the two with mean differences of -1.2 and -1.3 for the median ADC and IQR ADC values (Supplementary figure). The 95% limits of agreement were -8.3 to 6.0 for median ADC and -6.5 to 3.9 for IQR.

| **Supplementary table 2**: Summary of ADC estimates (mean and standard deviation, SD) using protocols A (Standard DWI) and B (non-accelerated DWI based on the research sequence) across all 20 lesions. | | | |
| --- | --- | --- | --- |
| ADC (x10^-5^ s/mm^2^) | Standard DWI | Non-accelerated research DWI | p-value of ADC estimates |
| Mean | 120.7 | 121.9 | 0.11 |
| SD | 41.7 | 41.5 |  |

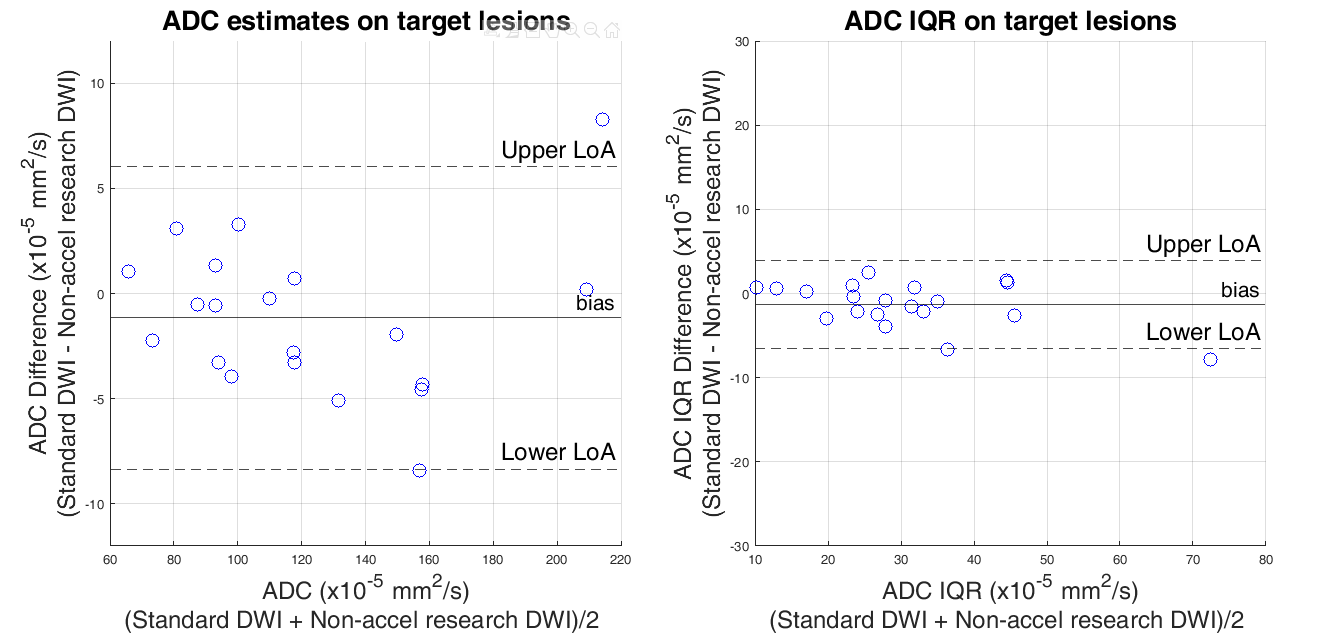

***Supplementary figure****: Bland–Altman plots of median (left) and IQR ADC (right) measurements demonstrating agreement between standard DWI and non-accelerated research DWI for the target bone lesions in Cohort 2. Two data points have nearly identical ADC IQR values and overlap, appearing as a single point on the right plot.*
